# Constructing genotype and phenotype network helps reveal disease heritability and phenome-wide association studies

**DOI:** 10.1101/2023.11.14.23297400

**Authors:** Xuewei Cao, Lirong Zhu, Xiaoyu Liang, Shuanglin Zhang, Qiuying Sha

**Author notes:** Corresponding author: QIUYING SHA, Department of Mathematical Sciences, Michigan Technological University, Houghton, Michigan 49931, USA. Both authors contributed equally (Xuewei Cao, Lirong Zhu).

## Abstract

Analyses of a bipartite Genotype and Phenotype Network (GPN), linking the genetic variants and phenotypes based on statistical associations, provide an integrative approach to elucidate the complexities of genetic relationships across diseases and identify pleiotropic loci. In this study, we first assess contributions to constructing a well-defined GPN with a clear representation of genetic associations by comparing the network properties with a random network, including connectivity, centrality, and community structure. Next, we construct network topology annotations of genetic variants that quantify the possibility of pleiotropy and apply stratified linkage disequilibrium (LD) score regression to 12 highly genetically correlated phenotypes to identify enriched annotations. The constructed network topology annotations are informative for disease heritability after conditioning on a broad set of functional annotations from the baseline-LD model. Finally, we extend our discussion to include an application of bipartite GPN in phenome-wide association studies (PheWAS). The community detection method can be used to obtain a priori grouping of phenotypes detected from GPN based on the shared genetic architecture, then jointly test the association between multiple phenotypes in each network module and one genetic variant to discover the cross-phenotype associations and pleiotropy. Significance thresholds for PheWAS are adjusted for multiple testing by applying the false discovery rate (FDR) control approach. Extensive simulation studies and analyses of 633 electronic health record (EHR)-derived phenotypes in the UK Biobank GWAS summary dataset reveal that most multiple phenotype association tests based on GPN can well-control FDR and identify more significant genetic variants compared with the tests based on UK Biobank categories.

## Introduction

The studies utilizing biological networks have proven to be successful in providing a comprehensive understanding of the complex relationships within the biological systems, such as gene regulatory networks^1; 2^, protein-protein interaction networks^3^, human disease networks^4^, et al. One of the commonly used biological networks is the bipartite network, which is defined as a network that consists of two distinct sets of nodes, with nodes in one set only connected to nodes in the other set and not within the same set. The human disease network usually describes the biological system as a bipartite network, where diseases and genes are represented as two distinct sets of nodes and disease nodes are only connected to their associated gene nodes. Rather than simply identifying the association between a genetic variant and a specific disease, the construction of a bipartite network can reveal the integrated molecular underpinnings of diseases^5^. Therefore, a bipartite network can be used to explore whether human diseases or complex traits and the corresponding genetic variants are related to each other at a higher level of cellular and organization^6; 7^. In addition, due to many complex diseases being affected by a shared set of pleiotropic variants, the construction of a bipartite network can also be used to determine the pathobiological relationship of one disease to other diseases^5^ and elucidate the complexities of genetic correlations across diseases^6^.

Over the past decade, genome-wide association studies (GWAS) have generated an impressive list of genetic variant and phenotype association pairs^8; 9^, which offer a great opportunity to establish a bipartite network connecting genetic variants and phenotypes, referred to as a genotype and phenotype network (GPN)^7^. GPN provides integrative analyses that allow for the characterization of complex relationships between genetic variants and phenotypes, which are reproducible and accurately represent biological relationships. Therefore, it has become increasingly important in recent years^10; 11^. Notably, a well-defined GPN is crucial as it provides a clear representation of the genetic association between genetic variants and phenotypes, including factors such as connectivity, centrality, and community structure. Meanwhile, the real-world biological network, including GPN, often exhibits a scale-free degree distribution^12; 13^, which means that a small number of nodes (genetic variants and phenotypes) have a much larger number of connections than the majority of nodes. In a random network, the nodes are connected randomly without any preferential attachment, resulting in a network with a relatively uniform degree distribution^14^. Therefore, comparing the degree distribution of a bipartite GPN to that of a random network can reveal important insights into the underlying mechanisms driving the construction of the network. Additionally, random networks can serve as a useful null model for testing the significance of network properties observed in the bipartite GPN.

The centralities of a bipartite GPN are one of the most important statistics to measure the importance of genetic variants (phenotypes) across phenotypes (genetic variants) based on the connectivity in the network^15^. The nodes with high centralities often act as hubs for information flow within the network^16^. For example, a genetic variant with high centrality accounting for all phenotypes is more likely to be a pleiotropic variant, as it is highly connected to multiple phenotypes in a bipartite GPN. Therefore, these centralities can be used to define the network topology annotations of genetic variants that quantify the possibility of a genetic variant being a pleiotropic variant. To study whether these network topology annotations are enriched for disease heritability, we apply stratified linkage disequilibrium (LD) score regression (S-LDSC)^17^ along with the leave-one-phenotype-out strategy to quantify the contribution of these annotations to disease heritability. We condition our analyses of the network topology annotations on the baseline-LD model, which includes a broad set of coding, conserved, regulatory, and LD-related functional annotations^18^. Additionally, in a bipartite GPN, a phenotype with a higher centrality accounting for all genetic variants is more likely to have a higher heritability, as it is connected to multiple genetic variants or with higher association evidence.

With the widespread availability of electronic health records (EHR) data, phenome-wide association studies (PheWAS) have been used to systematically examine the impact of one genetic variant across a broad range of phenotypes. Phenotypes in the whole phenome can be grouped by digitized codes (e.g., ICD-10 code) to represent the common clinical factors underlying the diseases. However, the taxonomy of digitized codes is based on their etiology rather than their genetic architecture, but applying the community detection method for GPN allows us to identify network modules that provide an integrative approach to understanding the complex genetic relationships across phenotypes^7^. A network module is loosely defined as a subnetwork with high local link density so that the phenotypes within a network module share more genetic architecture across all genetic variants than phenotypes outside the network module^19; 20^. Therefore, the network modules can serve as a priori grouping of phenotypes in PheWAS, allowing for jointly testing multiple phenotypes in each network module and a genetic variant to identify the cross-phenotype associations and pleiotropy. For multiple testing corrections, we apply a refined false discovery rate (FDR) control approach to obtain the significance thresholds for PheWAS.

## Material and Methods

In this section, we first describe our approach to constructing Genotype and Phenotype Networks (GPN) and defining the network topology annotations for genetic variants and phenotypes. The construction of GPN does not require access to individual-level genotype and phenotype data and only requires the marginal association evidence between each genetic variant and each phenotype (e.g., z-scores or estimated effect sizes from GWAS summary statistics). We first identify differences in denser representation and sparse representations of GPN with various sparsity approaches, then provide details of the implementation of constructed GPN, such as heritability enrichment of network topology annotations, estimation of the genetic correlation of multiple phenotypes, community detection of phenotypes, and phenome-wide association studies. **Figure 1** shows the workflow of this study.

**Figure 1.**
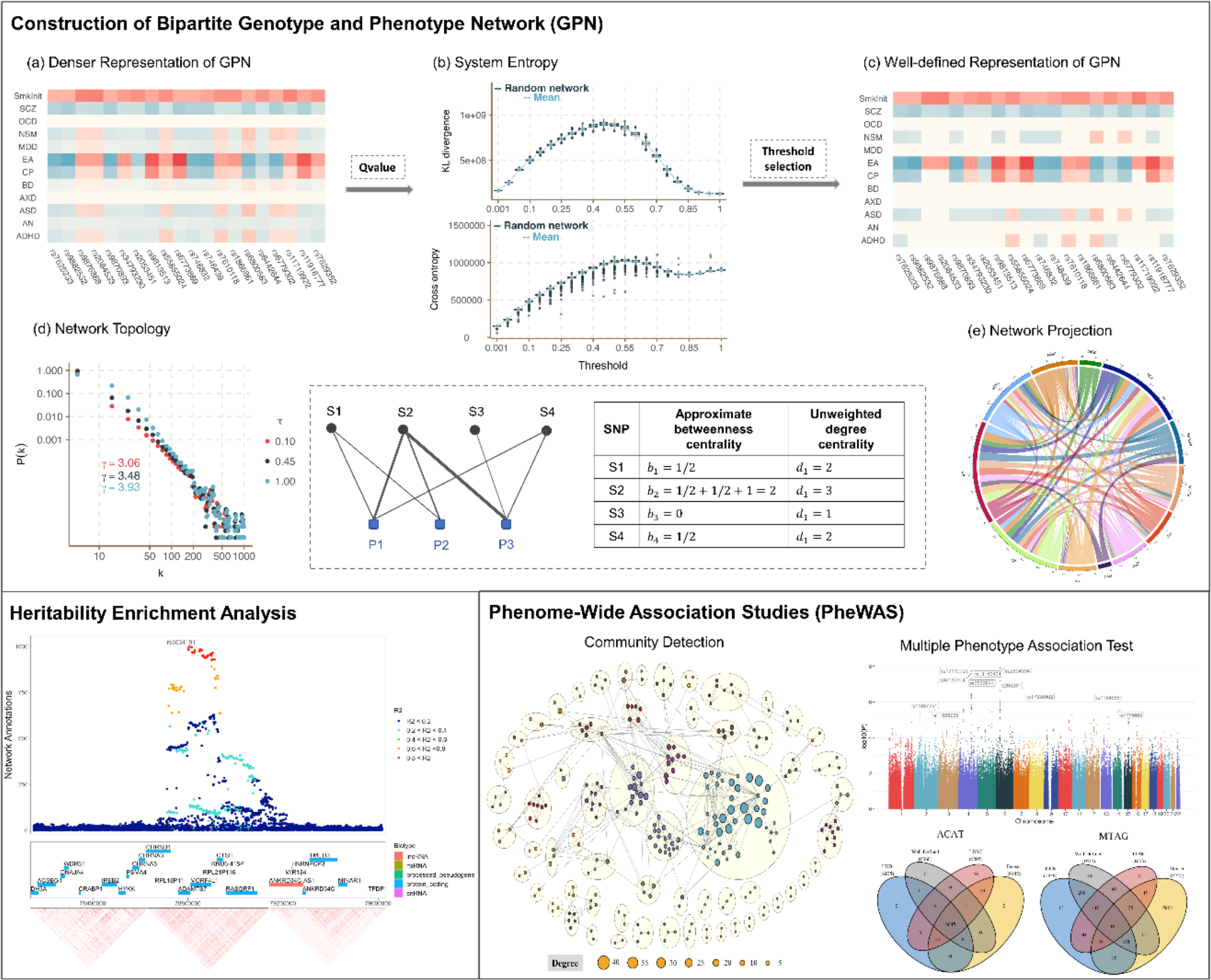
Graphical abstract. Construction of bipartite genotype and phenotype network (GPN) includes: (a) – (c) Construction of the denser and well-defined representations of GPN by comparing the network properties with the random networks, including connectivity, centrality, and system entropy; (d) The weighted degree distributions with different thresholds and the examples of two network topology annotations, approximate betweenness centrality and degree centrality, used in the heritability enrichment analysis; (e) The one-mode projection of GPN onto phenotypes that are linked through shared genetic architecture. Based on the constructed well-defined GPN, heritability enrichment analysis and phenome-wide association studies are introduced as two important applications of the constructed GPN.

### Bipartite genotype and phenotype networks construction

We consider GWAS summary statistical results from the same or different study cohorts with *K* phenotypic traits. Assume that the GWAS summary results for the *k*^*th*^ (*k* = 1, …, *K*) phenotype are calculated by testing the marginal association between a genetic variant and the *k*^*th*^ phenotype based on a sample with *N*_*k*_ unrelated individuals. Note that *N*_*k*_ = *N*_*l*_ (*k* ≠ *l*) if the GWAS summary statistics of the *k*^*th*^ phenotype and *l*^*th*^ phenotype are calculated from the same study cohort, otherwise, *N*_*k*_ ≠ *N*_*l*_. For simplicity, we assume the generalized linear regression^7^, 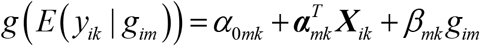, where *y*_*ik*_ is the *k*^*th*^ phenotype value and ***X***_*ik*_ is the vector of covariates, for example, used to account for population stratification in the study, for the *i*^*th*^ (1 ≤ *i* ≤ *N*_*k*_) individual and the *k*^*th*^ phenotype. Assuming that there are *M*_*k*_ genetic variants in the GWAS summary statistics for the *k*^*th*^ phenotype and *g*_*im*_ is the genotype of the *m*^*th*^ (1 ≤ *m* ≤ *M*_*k*_) genetic variant taking values from 0, 1, and 2 that counts the number of copies of the minor allele. Here, *g* (•) is either the identity link function for quantitative phenotypes or the logit link for binary phenotypes.

The GWAS summary results are calculated for testing the genetic association between the *k*^*th*^ phenotype and the *m*^*th*^ genetic variant under the null hypothesis *H* _0,*mk*_ : *β*_*mk*_ = 0. The commonly used Wald-type statistic is defined as 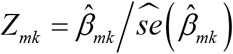 under the generalized linear regression model, where 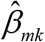 is the maximum likelihood estimation (MLE) of *β*_*mk*_ And 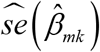 is its estimated standard error^21^. The p-value *p*_*mk*_ may also be calculated by assuming *Z*_*mk*_ ∼ *N* (0,1) in the GWAS summary results. In this study, we assume that only GWAS summary results (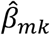 and *p*_*mk*_) are available.

Let *M* be the total number of unique SNPs included in the GWAS summary statistics for *K* phenotypes with the property of 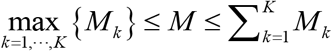. In particular, 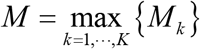 if and only if there is at least one GWAS summary data containing all unique genetic variants and 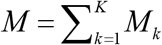 if and only if there are no variants included in different GWAS summary data. We can exclude the case 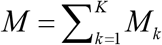 from our analyses since it rarely occurs in most GWAS summary datasets.

### Denser representation of GPN

Same as the network construction introduced by Gaynor et al.^11^, the denser representation of GPN allows us to capture the fact that we have no prior knowledge of precisely which genetic variants and phenotypes might have an association. Here, we construct the denser representation of GPN, an adjacency matrix that includes all the associations between genetics variants and phenotypes.

We first define a signed bipartite GPN, ***𝒢*** _*GPN*_ = (*Y,G, E*), where *Y* = {*Y*_1_,…,*Y*_*K*_ } and *G* ={*G*_1_,…,*G*_*M*_ } denote two disjoint and independent sets of phenotypes and genetic variants, and *E* denotes the set of edges in GPN. Similar to our previous work^7^, we denote **T** = (*T*_*mk*_) as an *M* × *K* adjacency matrix of the denser representation of GPN, where 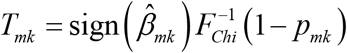 is the weight of the edge between the *m*^*th*^ genetic variant and the *k*^*th*^ phenotype. *F*_*Chi*_ (•) denotes the cumulative distribution function (CDF) of 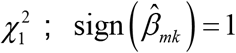 if 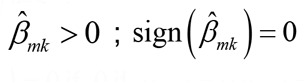; if 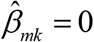; otherwise, sign 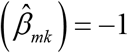. Note that |*T*_*mk*_| represents the strength of the association between the *m*^*th*^ genetic variant and *k*^*th*^ phenotype and sign 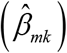 represents the direction of the association. The denser representation includes all associations and does not involve thresholding.

### Sparse representations of GPN

Given that even disease-associated genetic variants typically have a small effect size and are unlikely to exert their influence across the genome^11^, utilizing a sparsity-based approach makes biologically sense. Therefore, we introduce the false discovery rate (FDR) based sparse representations of GPN, in which the networks only include edges with associations meet a certain level of significance (i.e., p-value below a threshold) from the denser representation of GPN. Let 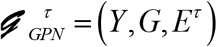 be a sparse representation of a bipartite GPN for a specific threshold *τ*, where *E*^*τ*^ denotes the set of edges in the sparse representation of 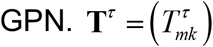 is an *M* × *K* adjacency matrix of GPN, where 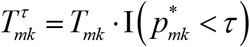 with *T*_*mk*_ from the denser representation of GPN. I (•) is an indicator function that takes value 1 when 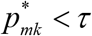, otherwise, it takes value 0. 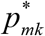 is a measure of the significance of genetic association between the *m*^*th*^ genetic variant and the *k*^*th*^ phenotype by correcting for multiple comparisons in each GWAS summary data. We use q-value^22; 23^ to define 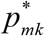 in our main analyses, but other adjustment methods for multiple comparisons can also be used, such as local FDR (LFDR)^24; 25^ and an adaptation of Benjamini-Hochberg (BH) FDR^26^. We use different Thresholds *τ* ∈[0,1], where *τ*= 1 represents the denser representation of GPN since all edges are included; *τ* = 0 represents the empty network with no edges between genetic variants and phenotypes.

#### Well-defined sparse representation of GPN

Selecting the appropriate threshold, *τ*, is very important in constructing GPN. The threshold is a sort of information filter, as decreasing *τ*, the resulting network will change from a denser network to a very sparse one. An overly dense network can be challenging in understanding and interpreting the most biologically informative interactions between genetic variants and phenotypes due to the abundance of information. Conversely, an excessively sparse network may lead to the loss of important information. The construction of a well-defined sparse representation of GPN can be presented to determine the optimal threshold 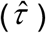 of 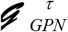, which can retain the key information about the interactions between genetic variants and phenotypes^27^. Therefore, we propose an approach to determine the optimal threshold by comparing the network properties with a corresponding random network, including connectivity, centrality, and community structure.

More specifically, we first calculate the network “connectance” for each 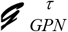, which is defined as the ratio of the number of edges in GPN to the total number of possible edges^28; 29^. Mathematically, it can be expressed as: *connectance*^*τ*^ = #{*E*^*τ*^ } (*M* × *K*), where #{•} is the counting measure, that is, #{*E*^*τ*^ } represents the number of edges included in 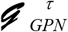. The degree of “connectance” in GPN can provide insight into the structure and functionality of the interactions between genetic variants and phenotypes. As decreasing *τ*, the resulting network will change from a dense network (*connectance*^*τ* =1^ ≈ 1) to a sparse one (*connectance*^*τ* =0^ = 0). For a specific *τ*, we then construct a corresponding random network by shuffling the edges of the original network 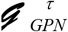. Let 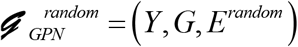 be the corresponding random network, where *conectance*^*τ*^ equals to *conectance*^*random*^. We also build an adjacency matrix **T**^*random*^ by keeping the same weights of the edges in *E*^*τ*^. Then, we compute the following network properties of 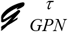 and 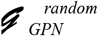, respectively.

*Weighted and unweighted degree*. The unweighted degree of a genetic variant (phenotype) in a bipartite GPN is defined as the number of edges across all phenotypes (genetic variants)^6^. The unweighted degree of the *m*^*th*^ genetic variant and the *k*^*th*^ phenotype are defined as 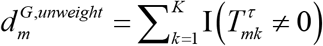 and 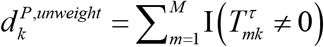, respectively. The weighted degree is reflecting the strength of the associations of edges, which are defined as 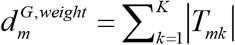 and 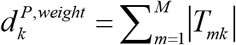.

*Kullback–Leibler (KL) divergence*. We define KL divergences^30; 31^ of degree of genetic variant and phenotypes between 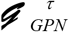 and 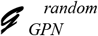 to determine the diversities between a bipartite GPN and a random bipartite network, which are given by

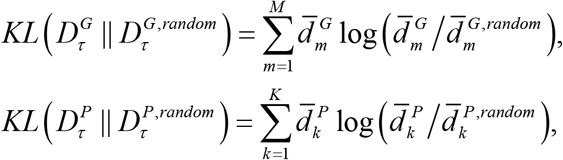

Where 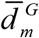 and 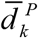 are the min-max standardized degree (either weighted or unweighted) which is defined as 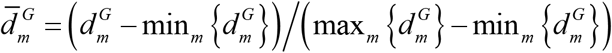 for the *m*^*th*^ genetic variant and 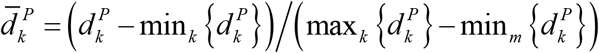 for the *k*^*th*^ phenotype. 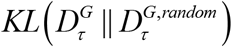 and 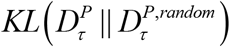 are used to measure the difference between degree distributions of genetic variants and phenotypes in 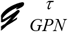 and 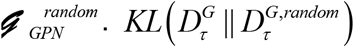 will equal 0 if the degree of genetic variants are the same in 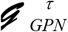 and 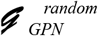 ; it will be negative if most degrees in 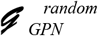 are greater than those in 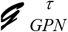 ; and it will be positive if most degrees in 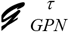 are greater than those in 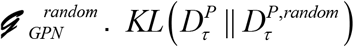 has same properties. We also define a global KL divergence of a bipartite network as 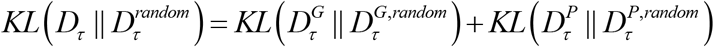.

Without loss of the generality, the optimal threshold *τ* should be selected by maximizing 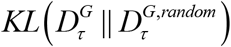 and 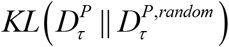. Meanwhile, considering the equivalent numbers and weights of edges in the original network and the corresponding random network, the greater the difference in network topologies between 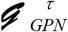 and 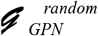, the more information 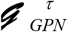 includes. To investigate the stability of the diversities, 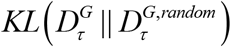 and 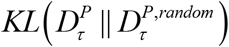, we construct 1,000 random networks for each 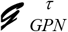. We thus can estimate the standard error of KL divergence and then obtain the stability by computing their 95% confidence intervals (CIs). We also evaluate two other network properties, degree entropy and cross entropy of degree (details in **Text S1**).

### Network topology annotations

For both denser and sparse representations of GPN, we constructed two probabilistic annotations based on the following network centralities. The centralities of a bipartite network are measuring the importance of genetic variants (phenotypes) across phenotypes (genetic variants) in the network. To simplify the notation, we use **T** to denote the adjacency matrix of GPN, which can be constructed by either a denser or sparse representation of GPN.

#### Degree centrality

In the context of bipartite GPN, a genetic variant with a high degree across phenotypes is more likely to be pleiotropic, owing to its strong connections with multiple phenotypes. Similarly, a phenotype with a high degree across genetic variants is more likely to have higher heritability and be associated with polygenic inheritance, as it is connected to multiple genetic variants or has stronger association evidence. The weighted degree of the *m*^*th*^ genetic variant or the *k*^*th*^ phenotype is defined as

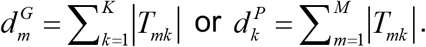

#### Approximate betweenness centrality

In a bipartite GPN, we define an approximate betweenness centrality of a genetic variant which can be used to measure its importance in connecting different phenotypes. A genetic variant with high approximate betweenness can be considered an important connector between phenotypes. The approximate betweenness centrality of the *m*^*th*^ genetic variant is defined as

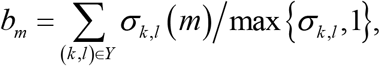

where *σ* _*k, l*_ is the number of shortest paths between the *k*^*th*^ phenotype and the *l*^*th*^ phenotype and *σ* _*k,l*_ (*m*) is the number of the shortest path between the *k*^*th*^ phenotype and the *l*^*th*^ phenotype that pass through the *m*^*th*^ genetic variant. Note that there are no direct edges between phenotypes in the bipartite GPN. Therefore, the shortest path *σ* _*k, l*_ is the number of genetic variants that are associated with both the *k* ^*th*^ phenotype and the *l*^*th*^ phenotype; the shortest path *σ* _*k, l*_ (*m*) only takes the value 0 or 1, where *σ* _*k, l*_ (*m*) = 1 if the *m*^*th*^ genetic variant is associated with both the *k*^*th*^ phenotype and the *l*^*th*^ phenotype, otherwise, *σ* _*k, l*_ (*m*) = 0.

### Heritability enrichment of network annotations

Note that the network topology annotations of genetic variants quantify the possibility of a genetic variant being a pleiotropic variant. To study whether these annotations are enriched for disease heritability of the highly correlated phenotype, we first perform a leave-one-phenotype-out (LOPO) approach to construct the network topology annotations. Then, we use stratified LD score regression (S-LDSC) to estimate the enrichment and the standardized effect size of the annotation^32; 33^.

#### Leave-one-phenotype-out (LOPO)

We consider *K* highly genetically correlated phenotypes. To simplify the notation, we use 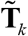 to denote the adjacency matrix of GPN by removing the *k*^*th*^ phenotype. 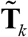 can be constructed by either denser or one of the sparse representations. Then, we use one of the network topology annotations based on the degree centrality and approximate centrality to assign the numeric value to each genetic variant for the evaluation of the *k*^*th*^ phenotype. Assigning a network topology annotation to each genetic variant is a way to quantify its potential for pleiotropy. The LOPO approach can assist in determining whether genetic variants have highly evidenced impacts on other *K* −1 phenotypes through pleiotropy and can also contribute to estimate the heritability of the *k*^*th*^ phenotype.

### Stratified LD score regression (S-LDSC)

S-LDSC is a method to assess the contribution of the annotation to disease heritability^32; 33^ conditional on other functional annotations. We use 86 functional annotations in the baseline-LD model (v2.1)^34^, including regulatory annotations (e.g., promoter, enhancer, histone marks, TF binding sites), LD-related annotations, et al. In this section, we omit the index *k* to simplify the notations. Let *a*_*mc*_ be the annotation value of the *m*^*th*^ genetic variant for the *c*^*th*^ annotation, where *m* = 1, …, *M*_*k*_ and *c* = 0, …, *C*. In particular, *a*_*m*0_ represent the network topology annotation of the *m*^*th*^ genetic variant constructed by the LOPO approach.

S-LDSC assumes that the per-SNP heritability or variance of the effect size of each genetic variant is given by 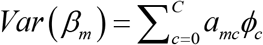, where *ϕ*_*c*_ is the per-SNP contribution of the *c*^*th*^ annotation to disease heritability. We can estimate *ϕ*_*c*_ using S-LDSC,

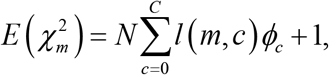

where 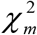 is the chi-square test statistic for testing the association between the *m*^*th*^ genetic variant and a phenotype in GWAS summary data, 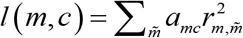 is the LD score of the *m*^*th*^ genetic variant to the *c*^*th*^ annotation, and 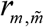 is the genotypic correlation between the *m*^*th*^ and the 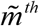 genetic variants.

We only focus on the network topology annotation *ϕ*_*0*_. As demonstrated by Finucane et al.^35^, *ϕ*_*0*_ will be positive if the network annotation increases per-SNP heritability, accounting for all other factors. Let *sd* (***a***_0_) be the standard deviation of the network topology annotation. The standardized effect size 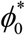 is defined by

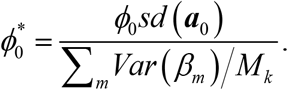

Note that 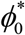 is defined as the proportionate change in per-SNP heritability associated with a one-standard-deviation increase in the network topology annotation conditioning on all other annotations^33^. The standard error on the estimate of 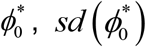, is computed using a block jackknife^32^. Then, we can compute the p-value to test if 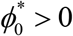 by assuming 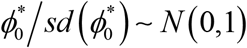.

We also calculate the enrichment of the network topology annotation, which is defined as the proportion of the heritability explained by genetic variants in the annotation divided by the proportion of genetic variants in the annotation.

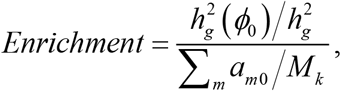

Where 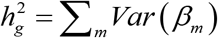 is the estimated heritability and 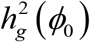 is the heritability captured by the network annotation. *Enrichment* > 1 represents the network annotation enriched for the disease heritability. Same as 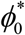, the significance for *Enrichment* is computed using a block jackknife^32^. The inclusion of the 86 functional annotations in the baseline-LD model can minimize the risk of bias in enrichment estimates and can also estimate the effect size *ϕ*_*0*_ conditional on the known functional annotations^32^.

### Community detection methods

Community detection methods are essential in comprehending the global and local structures of associations between genetic variants and phenotypes, and in shedding light on association connections that may not be easily visible in the network topology^15^. Calculating the projection of GPN onto phenotypes that are linked through shared genetic variants is a very important step in community detection. Let ***𝒢*** _*PPN*_ = (*Y, E*^*P*^) be the one-mode projection of GPN, called Phenotype and Phenotype Network (PPN), where *E*^*P*^ denotes the set of edges between phenotypes in PPN. Denote **W** = (*W*_*kl*_) as an *K* × *K* adjacency matrix of PPN, where *W*_*kl*_ is the weight of the edge between the *k*^*th*^ phenotype and the *l*^*th*^ phenotype. In this study, we perform community detection methods to partition *K* phenotypes into *L* disjoint network modules based on the adjacency matrix of PPN.

#### Community detection method for the denser representation of GPN

For the denser representation of GPN, one straightforward way to define the adjacency matrix **W** is to use the correlation of **T**, **W** = *cor* (**T**)^7^. The elements of **W** can be both positive and negative, implying that the PPN represented by the adjacency matrix of **W** is a signed network. Inspired by our previously proposed modularity-based community detection method^36^, we introduce a community detection method for the signed network in this study. Let 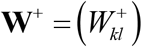 and 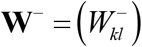 be adjacency matrices of the positive and negative weights, respectively, where 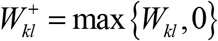 and 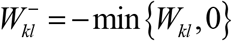 such that **W** = **W**^+^ − **W**^−^. First, we assume *K* phenotypes can be divided into *k*_0_ network modules using a hierarchical clustering method with similarity matrix **W** for *K*_*0*_ = 1, …, *K*. Let 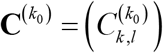 be a *K* × *K* connectivity matrix, where 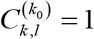 if the *k*^*th*^ phenotype and the *l*^*th*^ phenotype are in the same network module, otherwise, 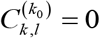. Then, we calculate the modularity of network with only positive weights, *W* ^+^, as 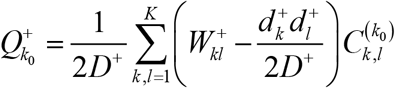 for each *k*_*0*_, where 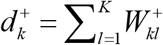 and 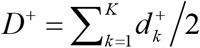 represent the degree of the *k*^*th*^ phenotype and overall degree of **W**^+^. Similarly, we calculate the modularity of **W**^−^ as 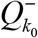. Therefore, we define the modularity for the signed network as 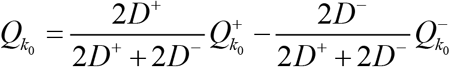. Note that a network’s modularity value indicates the density of connections within network modules and sparsity of connections between phenotypes in different models^15^. Then, we determine the optimal number of network modules as *L* = arg max{*Q*_1_,*Q*_2_, …,*Q*_*K*_ }.

#### Community detection method for the sparse representation of GPN

To eliminate the biases in projections caused by a large number of genetic variants that are unlikely to exert their influence across the whole genome^11^, we also provide a weighted projection approach by only focusing on the shared genetic variants between two phenotypes in the (well-defined) sparse representations of GPN, **T**^*sparse*^. Let 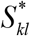 be the set of genetic variants that are connected with the *k*^*th*^ phenotype and the *l*^*th*^ phenotype. We define 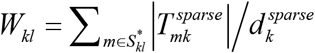 and 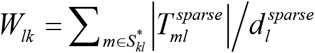, where 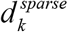 and 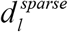 are the weighted degree of the *k*^*th*^ and the *l*^*th*^ phenotypes, respectively. More specifically, *W*_*kl*_ is a proportion of degree of the *k*^*th*^ phenotype explained by the shared associations between the *k*^*th*^ and the *l*^*th*^ phenotypes; similarly, *W*_*lk*_ is a proportion of degree of the *l*^*th*^ phenotype explained by the shared associations between the *k*^*th*^ and the *l*^*th*^ phenotypes. Therefore, *W*_*kl*_ ∆ *W*_*lk*_ indicates that the projected PPN is a directed network. If *W*_*kl*_ > *W*_*lk*_, the shared associations between the *k*^*th*^ and the *l*^*th*^ phenotypes are more important to the *k*^*th*^ phenotype than the *l*^*th*^ phenotype. In particular, *W*_*kl*_ = 1 if and only if the *k*^*th*^ phenotype only links with the genetic variants in 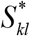. The modularity is easily generalized to both weighted and directed network, where the modularity based on LinkRank is given by^37; 38^: 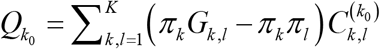. Let 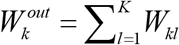be the out-degree of the *k*^*th*^ phenotype for a directed PPN. Then, *π* _1_, …, *π* _*K*_ is the PageRank vector indicating the probability of a phenotype being visited by a random surfer. 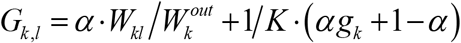 is the Google Matrix, where α is the damping parameter for PageRank^37^ (with probability 1−α random surfer jumps to a random phenotype) and 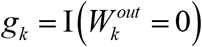 is an indicator of dangling phenotype. Same as the community detection method for the denser representation of GPN, we also determine the optimal number of network modules as *L* = arg max{*Q*_1_,*Q*_2_, …,*Q*_*K*_ }.

### Phenome-wide association studies (PheWAS)

The community detection method for PPN based on **W** has potential applications in PheWAS and multiple phenotype association studies. We extend our discussion to include the application of GPN in PheWAS. By using the community detection method of PPN, we can obtain a priori grouping of phenotypes and then jointly test the association between genetic variant and multiple phenotypes in each network module to discover the cross-phenotype associations and pleiotropy.

Assume that *K* is the total number of phenotypes in the whole phenome, which can be partitioned into *L* disjoint network modules by community detection. Let *K* = *K*_1_ + … + *K*_*L*_, where *K*_*l*_ is the number of phenotypes in the *l*^*th*^ network module. We apply four commonly used GWAS summary-based multiple phenotype association tests to identify the association between genetic variant and phenotypes in the *l*^*th*^ network module, including minP^39^, ACAT^40^, MTAG^41^, SHom^42^ (details in **Text S2**). Then, we refine our previous approach to evaluate FDR by thresholding the p-values obtained from the multiple phenotype association tests^43^. Let 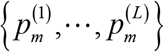 be a sequence of p-values for testing the association between phenotypes in each of the network modules and the *m*^*th*^ genetic variant. For a given nominal FDR level α ∈ (0,1), the optimal threshold for the *m*^*th*^ genetic variant is given by

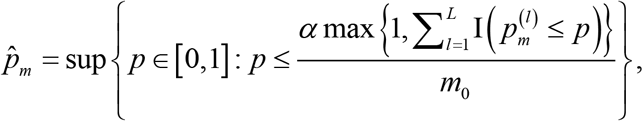

Where *m*_0_ is the number of network modules under the null hypothesis that phenotypes in the network module and the *m*^*th*^ genetic variant have no association. We use *m*_*0*_ = *L* − *m*_*1*_, where 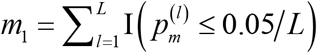 is the number of identified network modules that are associated with the *m*^*th*^ genetic variant based on the Bonferroni Correction.

### Empirical GWAS summary datasets

In our analyses, we consider two publicly available GWAS summary datasets to evaluate the performance of constructed bipartite GPN, heritability enrichment of network annotations, community detection methods, and the applications of PheWAS.

#### GWAS summary statistics for correlated phenotypes

To perform the heritability enrichment analysis of network annotations, we obtain publicly available GWAS summary data for 12 highly genetically correlated phenotypes in individuals of European ancestry, including attention deficit/hyperactivity disorder (ADHD), smoking initiation (SmkInit), autism spectrum disorder (ASD), neuroticism (NSM), anxiety disorder (AXD), major depressive disorder (MDD), obsessive-compulsive disorder (OCD), anorexia nervosa (AN), bipolar disorder (BD), schizophrenia (SCZ), educational attainment (EA), and cognitive performance (CP). The details of GWAS summary data for the 12 phenotypes are summarized in **Table S1**. As demonstrated by Zhang et al.^44^, the global genetic correlations among the 12 phenotypes estimated by their proposed SUPERGNOVA are ranging from -0.41 to 0.69. 51 out of 66 pairs of phenotypes have significant non-zero global genetic correlations (right upper triangle of **Table S2**). Meanwhile, they also reported the proportions of correlated regions between two phenotypes that are ranging from 0.11% to 93%. 46 pairs of phenotypes contain at least one significantly correlated region after Bonferroni correction (left lower triangle of **Table S2**). We only include the genetic variants in 22 autosomes.

#### GWAS summary statistics in the UK Biobank

The UK Biobank is a population-based cohort study with a wide variety of genetic and phenotypic information^45^. It recently released GWAS data on ∼ 500K individuals throughout the United Kingdom^46; 47^. For our study, we obtain the publicly available GWAS summary data for 633 EHR-derived phenotypes with main ICD10 diagnoses from Neale lab (Data availability). These GWAS summary data are calculated based on score tests on ∼337,000 unrelated individuals of British ancestry. We utilize the LD score regression (LDSC)^48^ on each of these 633 phenotypes, excluding 45 phenotypes from our analyses since the heritability estimators for them are out of bounds. There are 588 phenotypes across 1,096,648 genetic variants in autosomes in our analyses.

## Results

### Construction of GPNs for 12 genetically correlated phenotypes

We construct three bipartite GPNs for 12 genetically correlated phenotypes listed in **Table S1**, including a denser representation, an arbitrary sparse representation, and a well-defined representation. There are a total of 17,585,432 unique genetic variants from 12 GWAS summary datasets. The global genetic correlations and proportions of correlated regions among the 12 phenotypes estimated by SUPERGNOVA^44^ are shown in **Table S2**. We also perform LDSC^48^ to estimate phenotypic correlation (right upper triangle of **Table S3**) and genetic correlation (left lower triangle of **Table S3**) among the 12 phenotypes. Among a total of 66 pairs of phenotypes, 45 pairs of phenotypes have significant non-zero genetic correlations (p-values < 0.05/ 66 = 7.58×10^−4^). In particular, MDD has significant genetic correlations with all of the other 11 phenotypes, NSM has significant genetic correlations with 10 phenotypes except for BD, and SCZ and EA have significant genetic correlations with 10 other phenotypes but do not have significant genetic correlations with each other.

The denser representation of GPN is constructed without using any thresholds. Since the 12 GWAS summary datasets contain different numbers of the 17,585,432 unique genetic variants, the connectance of the denser representation of GPN is 0.5123 (**Figure S1(a)**). The well-defined sparse representation of GPN is constructed by comparing the network properties with the corresponding random networks. Since we have only 12 phenotypes in this analysis, we only consider the network properties for genetic variants of the constructed GPN and the corresponding random networks. For each *τ* ∈(0,1), we generate 1,000 corresponding random networks. **Figure 2 (a)** shows the comparisons of the KL divergence for genetic variants across 1,000 random networks. The KL divergence increases from 0 to a specific value of the threshold and then decreases from that value to 1, indicating that the difference between the original and random network reaches the maximum at the specific value. We also calculate the cross entropy of the weighted degree of genetic variants compared to the corresponding random network (**Figure 2 (b)**).

**Figure 2.**
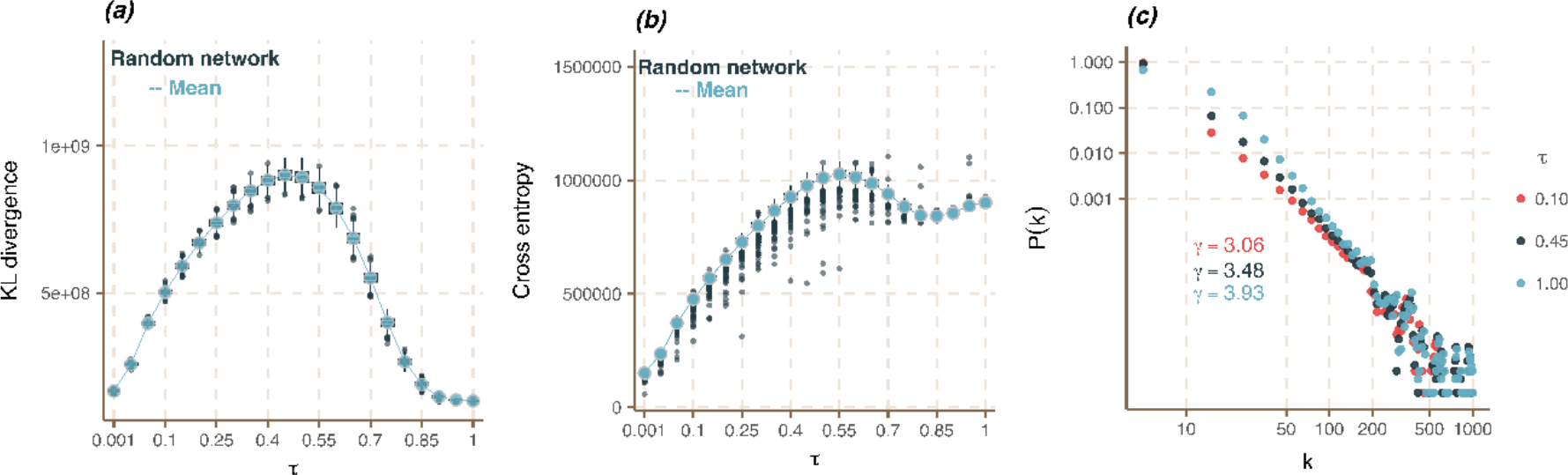
Network properties of the weighted bipartite GPNs for 12 genetically correlated phenotypes. (a) KL divergence for genetic variants. The blue line is the mean of KL divergencies across 1,000 random network comparisons. The boxplots show the scaled distributions of the KL divergence for each threshold. (b) Cross entropy for genetic variants. Blue lines are the means of cross entropy across 1,000 random network comparisons. The boxplot shows the scaled distribution of the cross entropy for each threshold. (c) Plot of the weighted degree distribution of genetic variants for three GPNs on the log-log scale, denser representation (*τ* = 1), well-defined sparse representation (*τ* = 0.45), and an arbitrary threshold sparse representation (*τ* = 0.1).

Note that the weighted degree of genetic variants in a corresponding random network becomes more different than the original one if the original network retains the key information about the interactions between genetic variants^27^. The network properties, KL divergence and cross entropy, will reach the maximum value at the most informative network. In our analysis, we prioritize choosing the optimal threshold with respect to KL divergence and then check the cross entropy and weighted degree entropy at that optimal threshold. The maximum mean of KL divergence equals 9.02 ×10^8^ at *τ* = 0.45, where the mean of cross entropy equals a larger value (9.83×10^5^) even though it does not reach the maximum value. Therefore, we constructed the well-defined sparse representation of GPN with *τ*= 0.45. This optimal threshold is much larger than the significant level for the association testing (e.g., *τ* = 0.05 for controlling FDR at the nominal level of 0.05). The optimal threshold in the construction of GPN does not represent the significant associations between genetic variants and phenotypes. It is only used to ensure that the constructed GPN is more informative than a random network.

As a comparison, we also construct an arbitrary sparse representation of GPN by using the threshold *τ* = 0.1. **Figure 2(c)** shows the weighted degree distribution of genetic variants for three GPNs, denser representation (*τ* = 1), well-defined sparse representation (*τ*= 0.45), and an arbitrary threshold sparse representation (*τ* = 0.1). We observe that the degree distributions of all three networks follow the power law with different scale parameters *γ*, indicating that a small number of genetic variants have a much larger number of connections than the majority of genetic variants. In particular, the degree of genetic variants in the denser representation of GPN is greater than those in a sparser GPN, resulting in the scale parameter increases with increasing the threshold *τ*.

We also calculate the network properties of the unweighted GPNs by comparing them with the corresponding random networks (**Figure S2**). Furthermore, the adjacency matrix of the projected PPN, **W** can be considered as the phenotypic correlation among 12 phenotypes based on the shared genetic architecture. **Figure S3** shows the comparisons of the adjacency matrix of PPN constructed by the denser and well-defined sparse representations of GPN with the genetic correlation matrix estimated by SUPERGNOVA^44^ (**Table S2**) and LDSC^48^ (**Table S3**).

### Heritability enrichment analysis of network annotations

For each of the three bipartite GPNs for the 12 phenotypes, we perform S-LDSC along with LOPO to evaluate whether the network topology annotations are enriched for disease heritability. We consider both degree centrality and betweenness centrality of genetic variants, conditioning on 86 functional annotations in the baseline-LD model (v2.1)^34^. These 86 existing functional annotations have been demonstrated to be highly informative by capturing functionality and LD-related features, thus, we evaluate the added value of our network topology annotations in capturing disease heritability, contributed by the pleiotropic variants with other genetically correlated phenotypes.

**Table 1** shows the heritability enrichment analysis results for degree centrality calculated from denser, arbitrary sparse, and well-defined sparse representations of GPN, respectively. From the LDSC results (**Table S3**), MDD has significant non-zero genetic correlations with all other 11 phenotypes. **Table 1** shows that the degree centrality annotation is significantly enriched for the heritability of phenotype MDD based on all of the three constructed GPNs (p-values<0.05/ 12 ≈0.0042). Specifically, the network topology annotation of each genetic variant quantifies its possibility for pleiotropy among other correlated phenotypes. After we use the LOPO approach to construct the network annotations of MDD, the significance enrichment indicates that the network annotation can contribute more information to disease heritability if it is computed based on other highly genetically correlated phenotypes. In particular, even though the arbitrary sparse representation of GPN (*τ* = 0.1) contains less information than the denser and well-defined GPN, the degree centrality annotation is still significantly enriched in heritability of MDD (p-value = 2.79×10^−5^) conditioned on the 86 functional annotations. Meanwhile, the degree annotation is also significantly enriched in heritability of CP (p-value = 2.76×10^−6^) and SCZ (p-value = 0.0021) for the arbitrary sparse representation of GPN. SCZ has significant non-zero genetic correlations with 9 phenotypes except for EA and CP (**Table S3**); CP has significant proportions of correlated regions with 9 phenotypes in which there are over 15% of correlated regions with 8 phenotypes (**Table S2**).

**Table 1.**
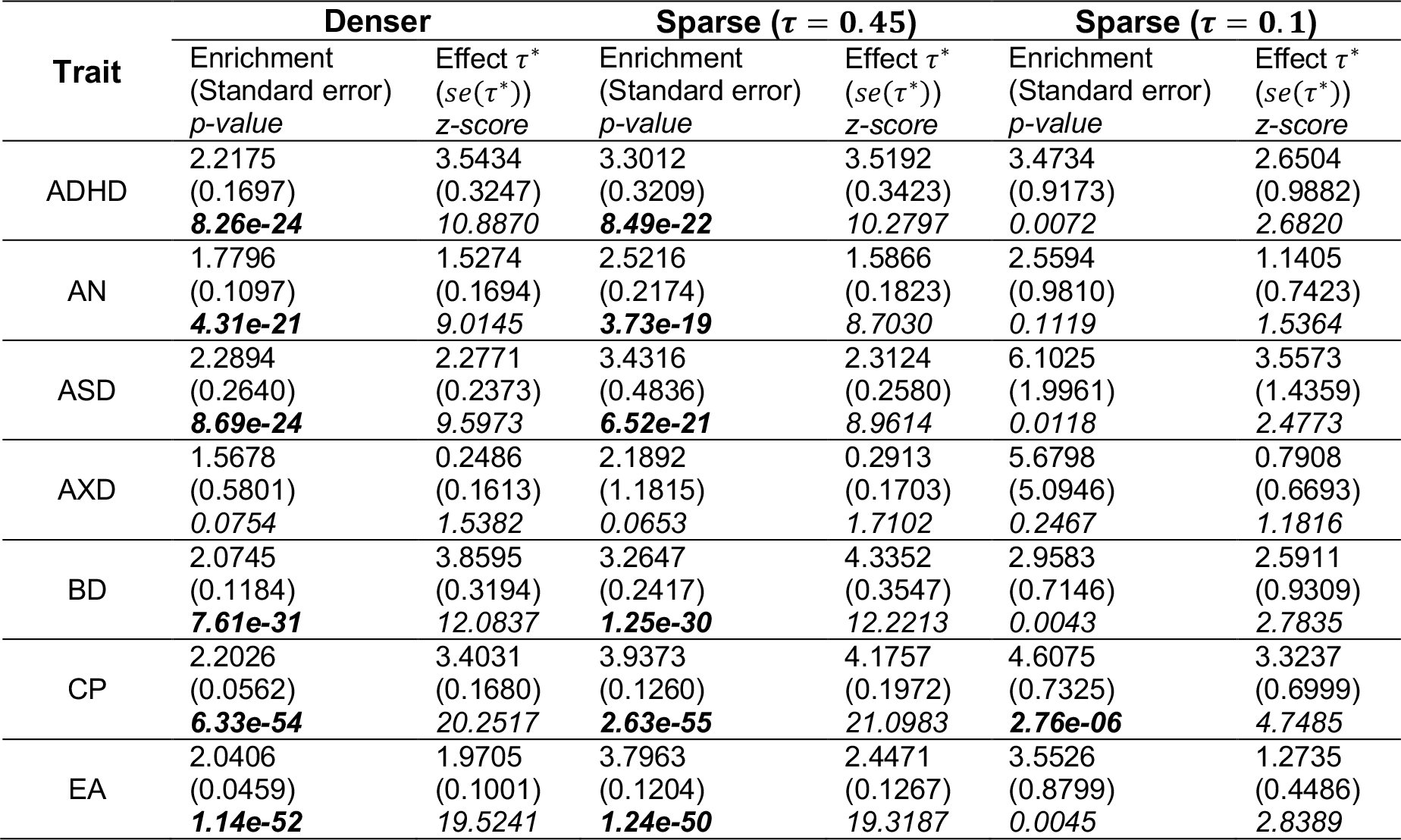
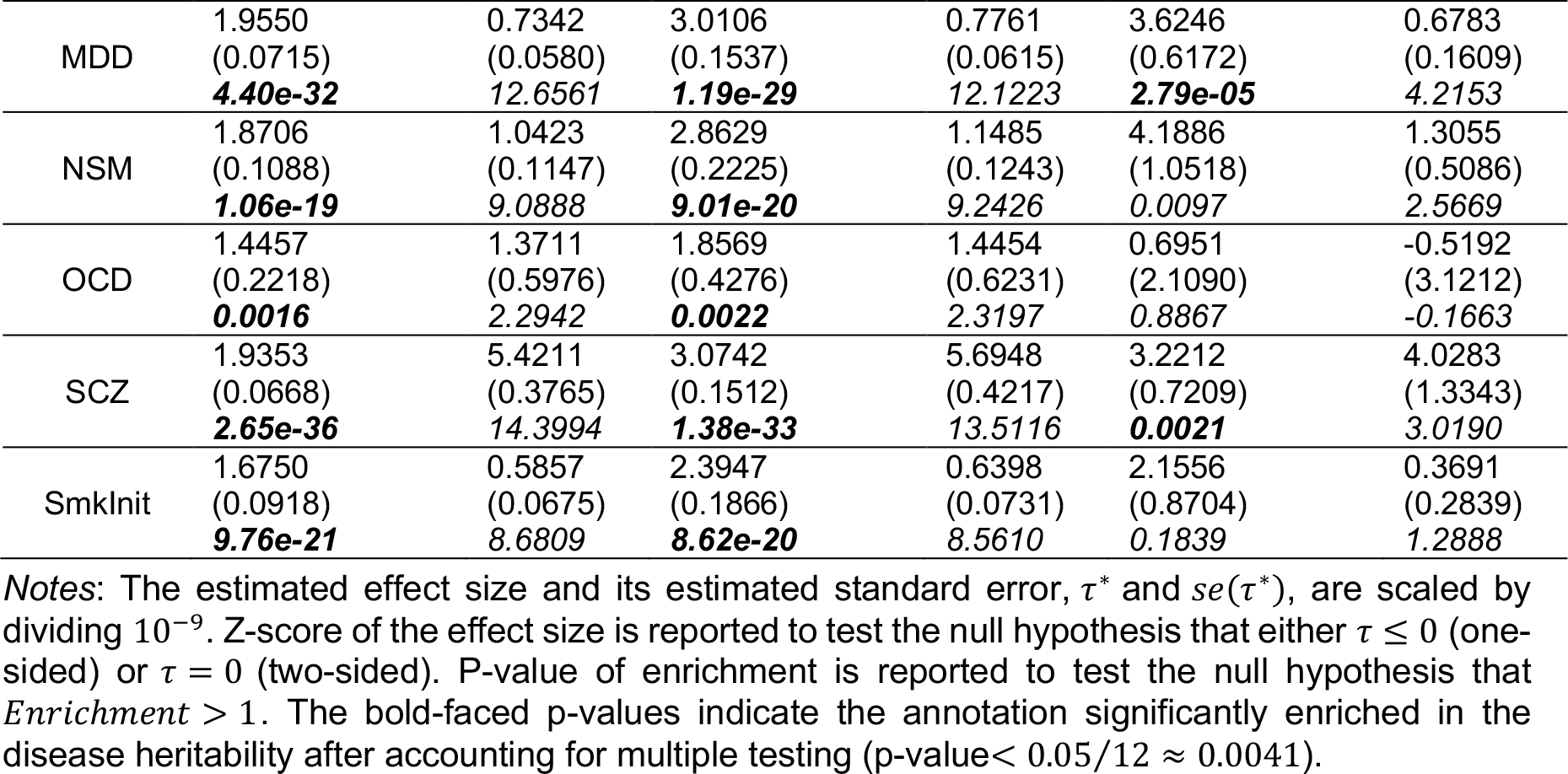
Heritability enrichment analyses of network topology annotation (degree centrality) based on denser and sparse representations of bipartite GPN for each of the 12 phenotypes.

The network annotation based on degree centrality obtained by the denser representation of a bipartite GPN includes the complete information for explaining the associations between phenotypes and genetic variants. It is significantly enriched to disease heritability of 11 out of 12 phenotypes as expected, except for AXD, with enrichment estimates ranging from 1.4457 (OCD with p-value = 0.0016) to 2.2894 (ASD with p-value = 8.69×10^24^). We identify the most significant enrichment of network annotations based on degree centrality for CP (Enrichment = 2.2026 with p-value = 6.33×10^−54^) and EA (Enrichment = 2.0406 with p-value = 1.14×10^−52^). These two phenotypes have a significant proportion of correlated regions, 93%, estimated by SUPERGNOVA^44^. **Figures S4(a) and S4(b)** show the QQ-plot of EA versus CP based on the weight of the denser and the well-defined sparse representations of GPN. Most of the genetic variants have similar weights for both EA and CP, lying in the diagonal line, but there exist some genetic variants that have the largest weights for only one phenotype. The same relationship between EA and CP is shown in the marginal associations from GWAS summary datasets (**Figures S4(c) and Figure S4(d)**).

The network topology annotations obtained by the well-defined sparse representation of GPN (*τ* = 0.45) perform similarly on the heritability enrichment compared to the denser representation of GPN. Even though some information is excluded from the well-defined GPN, the annotations obtained by the well-defined GPN contribute similar effects to disease heritability. **Table 1** and **Table S4** show that the annotations from both denser and well-defined sparse representations of GPN can significantly enrich disease heritability of the same phenotypes. However, the network topology annotations obtained by the arbitrary sparse representation of GPN (*τ*= 0.1) are not enriched to most disease heritability. We can conclude that a more informative network can be used to understand heritability rather than an arbitrary one with a smaller threshold. For example, if we use the significance level of the associations (e.g., *τ*= 0.1 or *τ*= 0.05) to construct a GPN, it may lose more information and key connections even though its edges represent the significant associations between genetic variants and phenotypes.

However, the network annotation based on approximate betweenness centrality performs differently on the heritability enrichment analysis than the annotation based on degree centrality. **Table S4** shows the heritability enrichment analysis results for betweenness centrality calculated from denser, arbitrary sparse, and well-defined sparse representations of GPN, respectively. We observe that the betweenness centrality calculated by the denser representation of GPN significantly enriches the disease heritability of only seven phenotypes, whereas the annotation calculated by the well-defined GPN can significantly enrich the heritability of 10 phenotypes. The strength of the associations between genetic variants and phenotypes is not considered in the betweenness centrality and the denser representation of GPN includes all edges. Therefore, the betweenness centrality of GPN is not an important feature that can be considered in the heritability enrichment analysis. Alternatively, it is an important network property for the sparse representation of GPN since only the edges with strong evidence of associations are included in the GPN. A genetic variant with high approximate betweenness can be considered an important connector between phenotypes. Therefore, the network annotations based on the approximate betweenness centrality calculated from the well-defined (*τ* = 0.45) and the arbitrary (*τ* = 0.1) sparse representation of GPN are significantly enriched to 10 phenotypes’ heritability. Meanwhile, the network annotation calculated by a well-defined GPN has stronger evidence than that calculated by the arbitrary one.

According to heritability enrichment results, we observe that network annotations are not enriched to the disease heritability of AXD and OCD. **Figure S5** shows the heatmap of edge weights in the well-defined sparse representation of GPN for the top 100 and the top 1000 genetic variants with the highest degree of centrality, respectively. We observe that these top genetic variants have smaller weights on AXD and OCD, which means that the genetic variants with the highest degree of centrality are not associated with AXD and OCD. Therefore, the network annotation is not enriched to their heritability. In particular, there are no edges between OCD and genetic variants if the threshold is smaller than 0.4.

### Construction of GPNs for 588 EHR-derived phenotypes in the UK Biobank

For a total of 1,096,648 genetic variants and 588 EHR-derived phenotypes with main ICD10 diagnoses after preprocessing, we construct two bipartite GPNs including a denser representation and the well-defined sparse representation. Different from the previous 12 GWAS summary datasets obtained from different studies, GWAS summary datasets of these 588 phenotypes are calculated based on score tests on the same ∼337,000 unrelated individuals of British ancestry.

Therefore, the connectance of the denser representation of GPN equals 1, that is, all genetic variants link with all phenotypes with strength of the associations (**Figure S1(b)**).

We consider the network properties for both genetic variants and phenotypes of constructed GPN and the corresponding random networks. For each *τ* ∈(0,1), we generate 1,000 corresponding random networks. **Figures 3(a) and 3(b)** show the KL divergence for genetic variants and phenotypes across 1,000 random network comparisons, respectively. The KL divergence increases from 0 to a specific value of the threshold and then decreases from that value to 1, indicating that the difference between the original and random network reaches the maximum at the specific value. We also calculate the cross entropy and degree entropy of the weighted degree of genetic variants compared to the corresponding random network (**Figure S6**). The maximum mean of KL divergence equals 1.14×10^8^ at *τ* = 0.6, where the mean of cross entropy equals 3.90 ×10^4^ with the largest standard error (17.08) compared with other thresholds. Therefore, we constructed the well-defined sparse representation of GPN with *τ*= 0.6. We also compare degree distributions of the well-defined network with a denser representation (*τ* = 0.8) and two arbitrary threshold sparse representations (*τ* = 0.2 and *τ* = 0.4) of GPN. Similar to the constructed GPN of 12 genetically correlated phenotypes, the degree distributions of all four networks follow the power law with different scale parameters *γ*, indicating that a small number of genetic variants have a much larger number of connections than the majority of genetic variants. In particular, the degree of genetic variants in the denser representation of GPN is greater than those in the sparser GPNs, resulting in the scale parameter increases with increasing the threshold *τ*. Meanwhile, we calculate the network properties of the unweighted GPNs by comparing them with the corresponding random networks (**Figure S7**).

**Figure 3.**
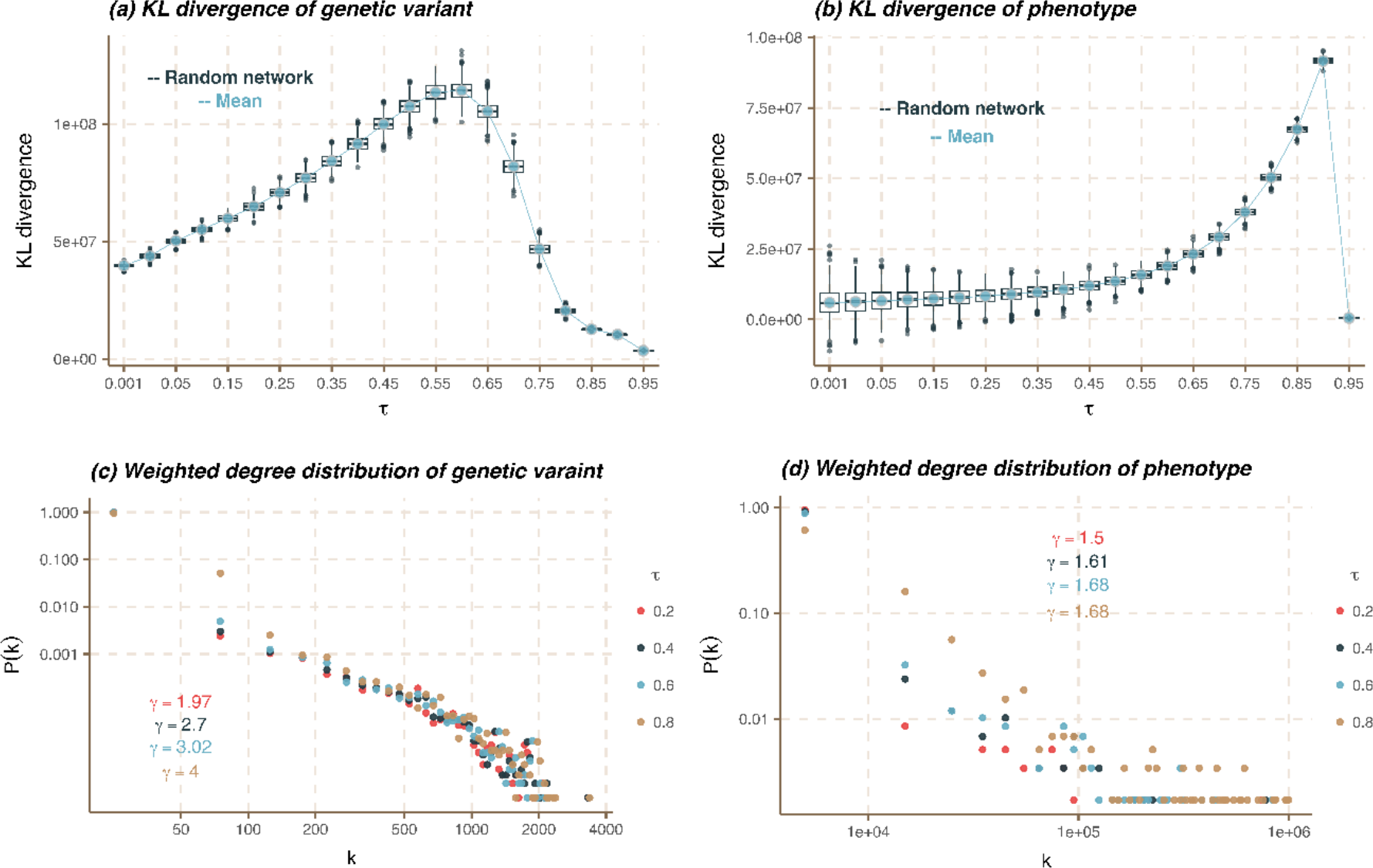
Network properties of the bipartite GPNs for 588 EHR-derived phenotypes in the UK Biobanks. (a) and (b) KL divergence for genetic variants and phenotypes. The blue line is the mean of KL divergencies across 1,000 random network comparisons. The boxplots show the scaled distribution of KL divergence for each threshold. (c) and (d) Weighted degree distribution of genetic variants and phenotypes for four GPNs on log-log scale, denser representation (*τ* = 0.8), well-defined sparse representation (*τ* = 0.6), and two arbitrary threshold sparse representations (*τ* = 0.2 and *τ* = 0.4).

We calculate three network topology annotations of genetic variants in the constructed GPNs with *τ* = 0.2, 0.4, 0.6, 0.8, including weighted degree centrality, unweighted degree centrality, and approximate betweenness centrality (**Figure S8 and S9**). **Figure S8** illustrates the relationship between the approximate betweenness centrality of genetic variants and the weighted degree centrality of genetic variants. The top five genetic variants with the highest degree and centrality are marked, respectively. These variants have mostly been associated with multiple phenotypes in the GWAS Catalog, and they overlap considerably under different parameter τ. Using the optimal parameter (*τ* = 0.6), we have summarized the number of significantly associated phenotypes in **Table S5**. Additionally, the top five genetic variants with the highest weighted degree centrality are almost entirely located in the same LD blocks. However, the top five genetic variants with the highest approximate betweenness centrality are associated with multiple phenotypes and display a pleiotropic effect among them. Similarly, we also compare the relationship between the approximate betweenness centrality of genetic variants and the unweighted degree centrality of genetic variants (**Figure S9**). **Table S6** shows the top five genetic variants with highest unweighted degree and approximate centralities.

### Community detection for phenotypes

For the denser representation of GPN, we construct the one-mode projected PPN by taking the correlation of the adjacency matrix of GPN. After applying the modularity-based community detection method to the signed PPN, we partition 588 EHR-derived phenotypes into 132 disjoint network modules. The number of phenotypes in each network module ranges from 1 to 87. For the well-defined sparse representation of GPN, we also construct a directed PPN by only focusing on the shared genetic variants between two phenotypes. In the sparse representation of GPN, phenotypes link with multiple genetic variants, but different phenotypes may not share a link with the same genetic variants. That is, we define the adjacency matrix for the *k* ^*th*^ phenotype as *W*_*kl*_ = 0 for all *l* = 1, …, *K* if the *k*^*th*^ phenotype does not share the same genetic variants with other phenotypes. Therefore, we first isolate 125 phenotypes without sharing any genetic variants with other phenotypes as 125 network modules for a single phenotype. Then, we partition the remaining 463 phenotypes into 71 network modules using the community detection method introduced in method. The number of phenotypes in the 71 network modules ranges from 2 to 37, and there are a total of 196 network modules. For comparison, we also apply our proposed community detection method based on the denser representation of GPN to LDSC phenotypic correlation. 588 phenotypes are divided into 114 categories with the number of phenotypes ranging from 2 to 82.

### PheWAS for 588 EHR-derived phenotypes in the UK Biobank

In PheWAS, a priori grouping (network module) of phenotypes in whole phenome can be obtained by the community detection of PPN. For each network module, we jointly test the phenotypes within this module and a genetic variant to discover the cross-phenotype associations and potential pleiotropy. In this study, we perform four most commonly used GWAS summary-based multiple phenotype association tests to identify the association between phenotype in each network module and each of genetic variants, including minP^39^, ACAT^40^, MTAG^41^, and SHom^42^ (details in **Text S2**). Then, we use the refined FDR controlling approach to evaluate FDR by thresholding the p-values obtained from the multiple phenotype association tests.

#### Simulation studies

We first conduct extensive simulation studies to evaluate whether these four multiple phenotype association tests used in our study can well-control FDR. We consider two simulation settings: 500 phenotypes with 50 phenotypic categories and 1,000 phenotypes with 100 phenotypic categories (details in **Text S3**). We assume that only the phenotypes within the same phenotypic category are correlated with each other. Similar to Lee et al.^49^, we consider two scenarios of correlations among phenotypes within the same category: 1) same correlation between each pair of phenotypes (SAME); 2) different correlation between each pair of phenotypes that is generated by using an autoregressive (AR(1)) model. **Table S7** and **Table S8** show the average FDR in the simulation studies for 500 phenotypes and 1,000 phenotypes, respectively. FDR is evaluated using 10 Monte-Carlo (MC) runs, equivalent to 1,000 replications at a nominal FDR level of 5% (**Text S3**). The 95% confidence interval (CI) is (0.0365, 0.0635). Note that we directly generate z-scores instead of effect sizes of genetic variants on phenotypes without considering LD, therefore, we do not consider MTAG in our simulation studies. The correlations among phenotypes are estimated by the method introduced in Kim et al.^39^. We observe that minP cannot control FDR in all scenarios but ACAT, and SHom can well control FDR as expected.

#### PheWAS based on 165 UK Biobank level 1 categories

As benchmarked categories, 588 EHR-derived phenotypes are grouped into 165 UK Biobank level 1 categories defined in data-field 41202 (https://biobank.ndph.ox.ac.uk/showcase/field.cgi?id=41202). The number of phenotypes in each category ranges from 1 to 20: there are 43 categories containing only one phenotype; 35 and 31 categories contain 2 and 3 phenotypes, respectively; only 7 categories contain more than 10 phenotypes. In our real data analyses, we only apply three multiple phenotype association tests (ACAT, SHom, and MTAG) to test the association between phenotypes in each category and each genetic variant. minP is not considered here since it cannot control FDR evaluated in our simulation studies. We use the commonly used genome-wide nominal FDR level 5×10^−8^. After applying our refined FDR controlling approach for the tests of each genetic variant, ACAT can identify 6,105 genetic variants associated with at least one category. We observe that most genetic variants are associated with only one category. SHom can identify 2,701 genetic variants and MTAG can identify 2,980 genetic variants (**Figure 4**).

**Figure 4.**
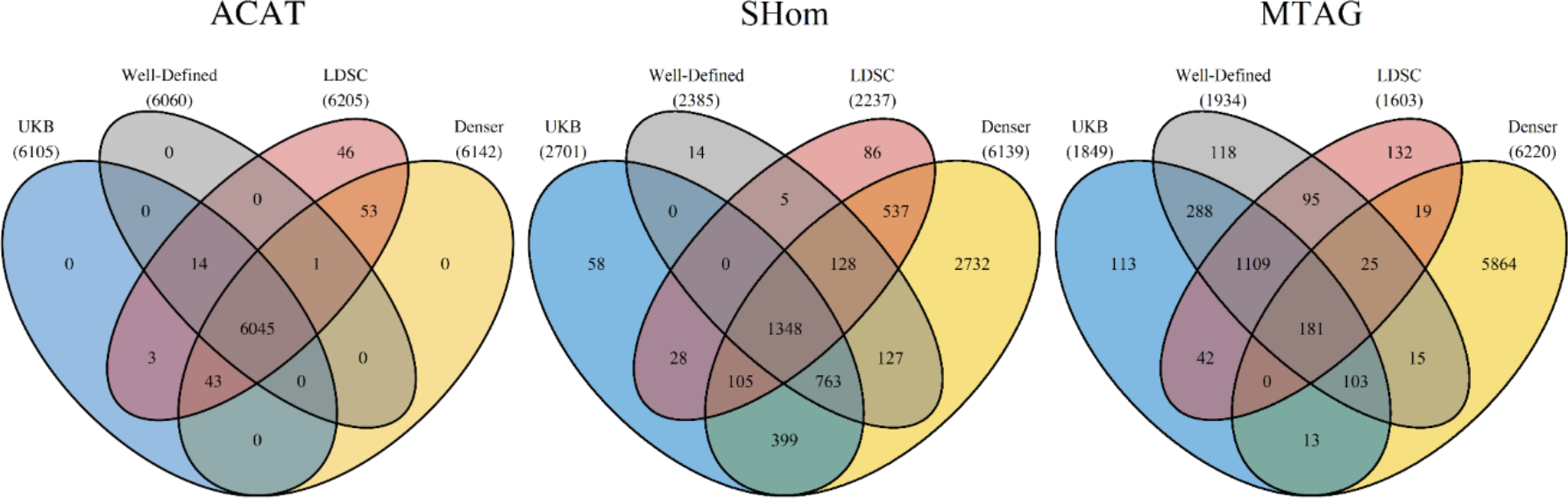
Venn plots for genetic variants identified by three multiple phenotype association tests based on different phenotypic categories and network modules.

#### PheWAS based on 114 phenotypic categories from LDSC

As a comparison, there are 114 phenotypic categories of the 588 EHR-derived phenotypes detected from the phenotypic correlation estimated by LDSC. We also apply three multiple phenotype association tests to 114 categories. ACAT identifies 6,205 genetic variants, SHom identifies 2,237 genetic variants, and MTAG identifies 1,603 genetic variants. Compared with the association tests based on the phenotypic categories in the UK Biobank, ACAT based on the LDSC can identify all of the 6,105 genetic variants identified by ACAT based on the UK Biobank (**Figure 4**). Meanwhile, 100 genetic variants are uniquely identified by ACAT based on the LDSC. **Figure S10** shows the heatmap of -log10(p-value) from GWAS summary datasets of these 100 genetic variants. We observe that all of these 100 genetic variants are significantly associated with at least one phenotype at the GWAS significance level 5×10^−8^. According to results from SHom and MTAG, tests based on the UK Biobank identify more genetic variants than the tests based on the LDSC.

#### PheWAS based on 132 network modules from the denser representation of GPN

Based on the denser representations of GPN, 588 EHR-derived phenotypes are partitioned into 132 disjoint network modules According to these 132 network modules, ACAT can identify 6,142 genetic variants associated with at least one network module and SHom can identify 6,139 genetic variants. In the application of MTAG, it is time-consuming and out of memory for one network module with 87 phenotypes. Therefore, we perform MTAG on the other 131 network modules and MTAG identifies 6,220 genetic variants. **Figure 4** shows the Venn plot for genetic variants identified by three multiple phenotype association tests based on different phenotypic categories and network modules. Based on the network modules detected from the denser representation of GPN, all three methods (ACAT, SHom, and MTAG) can identify ∼6,000 genetic variants associated with at least one network module.

#### PheWAS based on 196 network modules from the well-defined representation of GPN

Based on the well-defined representation of GPN, 588 EHR-derived phenotypes are partitioned into 196 network modules. According to the 196 network modules, ACAT can identify 6,060 genetic variants associated with at least one network module; SHom can identify 2,385 genetic variants; and MTAG can identify 1,934 genetic variants. From ACAT results, 6,060 genetic variants are identified by ACAT based on at least two other grouping of phenotypes, even if it identifies a smaller number of genetic variants. According to results from SHom and MTAG, tests based on the network modules detected from well-defined GPN identify more genetic variants than the tests based on the LDSC and the UK Biobank, but they identify fewer genetic variants than the tests based on the network modules detected from denser GPN.

## Discussion

In this paper, we conduct a comprehensive analysis to build the GPNs, which can be a routine procedure in post-GWAS investigations. Owing to increasingly accessible to GWAS summary statistics, the construction of GPN only requires the marginal association evidence between each genetic variant and each phenotype in GWAS summary data instead of individual-level genotypes and phenotypes data. The denser representation of the bipartite GPN can be directly constructed by linking all genetic variants and phenotypes in GWAS summary datasets. Although a denser representation of bipartite GPN contains information on all pairwise associations between genetic variants and phenotypes, pruning the network is both biologically meaningful and computationally efficient^11^. However, the thresholding approach used to prune is significantly influenced by the network size (connectance). To address this issue, we propose to construct a well-defined GPN with a clear representation of genetic associations by comparing the network properties with a random network, including connectivity, centrality, and community structure. Our findings indicate that a well-defined network with an optimal threshold can preserve crucial information on the associations between genetic variants and phenotypes.

Based on the construction of the denser and well-defined representation of bipartite GPNs, we further propose two network topology annotations based on the degree centrality and the approximate betweenness centrality. Both of the annotations can be used to quantify the possibility of pleiotropy for genetic variants. We highlight one of our significant discoveries that link pleiotropy and disease heritability through the utilization of heritability enrichment analysis using the stratified LD score regression. We analyze 12 genetically correlated phenotypes to show that the genetic variants with high degree centrality and approximate betweenness centrality are enriched for disease heritability conditioning on known functional annotations from the baseline LD model. First, in the analyses of the degree centrality based on the denser and the well-defined GPNs, we identify 10 phenotypes with significant heritability enrichment after using the LOPO approach. The significant enrichment indicates that the degree annotation can contribute more information to disease heritability if it is computed based on other highly genetically correlated phenotypes. We also observe that the denser GPN provides more information in the degree centrality as the degree centrality contains the strength of marginal association evidence. Second, we determine that network annotation based on the approximate betweenness centrality calculated from the well-defined GPN is strongly enriched for disease heritability. However, the disease heritability of some phenotypes is fully explained by annotations from the baseline-LD model in the analysis of the approximate betweenness centrality calculated from the denser GPN.

Construction of the bipartite GPNs also has important implications for the PheWAS. In particular, detecting the network modules of phenotypes from the constructed GPN is essential in understanding the global and local structures of associations between genetic variants and phenotypes, and in shedding light on association connections that may not be easily visible in the network topology. The detected network modules can be used as a priori grouping of phenotypes in PheWAS, then jointly testing of multiple phenotypes in each network module and one genetic variant can be performed to discover the cross-phenotype associations and pleiotropy. Significance thresholds for PheWAS are adjusted for multiple testing by applying the false discovery rate (FDR) control approach. First, we discover that the three multiple phenotype association tests (ACAT, SHom, and MTAG) applied in this study can well-control FDR as demonstrated by extensive simulation studies. Second, we analyze 633 EHR-derived phenotypes in the UK Biobank GWAS summary datasets. Based on the network modules detected from the denser representation of GPN, all three tests can identify more genetic variants associated with at least one network module (∼6,000 genetic variants) compared with the tests based on the UK Biobank, LDSC, and well-defined GPN.

There are some limitations to the work presented here. First, genetic effects can be heterogenous across phenotypes and studies based on different GWAS summary statistics^50; 51^ due to different sample sizes, genetic architectures, and quality of the genotyping and phenotyping data, et al. In our current analyses, we ignore the influence of different sample sizes for different GWAS summary statistics in the construction of GPN. However, larger sample sizes are typically associated with smaller standard errors and more precise effect size estimates, which can help to reduce bias and increase the stability of effect size estimates. To construct a GPN with stable evidence of the associations in the edges, we suggest that the sample sizes used to calculate the GWAS summary results in each study are sufficiently large (e.g., *N*_*k*_ > 10, 000). Second, we use the marginal association between each genetic variant and each phenotype to define the edge of GPN. The challenge in validating our proposed construction of GPNs is that there is no source of “ground truth” of GWAS. There may exist spurious associations between multiple genetic variants and a phenotype due to LD^9^. For example, a genetic variant in high LD with a true causal variant may be detected instead of the causal variant itself. However, several powerful fine-mapping and colocalization approaches have been developed to leverage information on LD to identify the putative causal variants in a specific genomic region^52-54^, which provides a great opportunity to construct a more informative GPN for future studies. Third, we do not consider the functional relationships between genetic variants and phenotypes. Filtering candidate (functional) regions based on the strength of gene-based associations may reduce multiple testing burdens and consequently improve statistical power in the construction of GPN. For example, transcriptome-wide association studies can combine genetic and transcriptomic data in a specific tissue to identify functional variants and genomic regions, which provide insights into biological pathways^55^.

## Data Availability

All data produced in the present work are contained in the manuscript

## Acknowledgments

Part of this research has been conducted using the UK Biobank resource under application number 102999 and the NHGRI-EBI GWAS Catalog. The work was in part funded by the Portage Health Foundation Graduate Assistantship, and Michigan Technological University Graduate Dean Awards. High-Performance Computing Shared Facility (Superior) at Michigan Technological University was used in obtaining results presented in this publication.

## Author contributions

Formal analysis and Methodology: XC, LZ, XL, SZ, and QS; Data curation and Visualization: XC and LZ; Writing original draft: XC, LZ, and QS; Writing review and editing: XC, LZ, XL, SZ, and QS.

## Web resources

### Data

GWAS summary statistics for 12 highly correlated phenotypes can be downloaded from the corresponding consortium websites reported in Zhang et al.^44^.

GWAS summary statistics for 633 EHR-derived phenotypes with main ICD10 diagnoses can be found from Neale lab: http://www.nealelab.is/blog/2017/7/19/rapid-gwas-of-thousands-of-phenotypes-for-337000-samples-in-the-uk-biobank.

### Software

PLINK version 1.9 can be downloaded from https://www.cog-genomics.org/plink/1.9/ ^56^.

LDSC: the command line tool for estimating heritability and genetic correlation from GWAS summary statistics can be downloaded from https://github.com/bulik/ldsc ^57^.

Cytoscape: an open-source software platform for visualizing complex networks which can be accessed via https://cytoscape.org/ ^58^.

## Data and code availability

This study does not generate new data. The codes generated during this study are available at a public repository https://github.com/xueweic/GPN.

